# Large vessel occlusions of milder severity show better collaterals and reduced harm from thrombectomy transfer delays

**DOI:** 10.1101/2023.06.05.23291002

**Authors:** Hannah Rowling, Dominic Italiano, Leonid Churilov, Logesh Palanikumar, Jackson Harvey, Timothy Kleinig, Mark Parsons, Peter Mitchell, Stephen Davis, Nyika Kruyt, Bruce Campbell, Henry Zhao

## Abstract

**Background and Purpose:** Patients with large vessel occlusion (LVO) stroke of milder baseline clinical severity are often missed by pre-hospital severity-based triage tools. We examined whether these patients had differences in 1) markers of collateral circulation and 2) the relative harm of transfer delays on outcome, compared to patients with clinically more severe baseline deficits.

**Methods:** Registry data from two large Australian thrombectomy centers was used to identify all directly presenting and transferred LVO patients undergoing endovascular thrombectomy, divided into those with milder baseline deficits (NIHSS <10) and severe baseline deficits (NIHSS ≥10). Baseline CT-perfusion markers of collateral circulation and the association between transfer status and functional outcome (using the 90-day modified Rankin Scale) were compared.

**Results:** Of 1210 included LVO patients, 273 (22.6%) had milder severity. Milder LVO patients showed smaller median ischemic core volumes (12.6 [IQR 0.0-17.9] vs 27.5 [IQR 6.5-37.1] mL, p<0.001)), higher median perfusion mismatch ratio (148.3 [IQR 5.0-500.0] vs 66.2 [IQR 3.6-17.2], p<0.001) and lower median hypoperfusion intensity ratio (0.25 [IQR 0.18-0.38] vs 0.40 [IQR 0.22-0.57], p<0.001). Transferred patients had a lower odd of excellent outcome than primary presenters, when deficits were severe deficits (aOR 0.759 [95% CI 0.576-0.999]) but not when deficits were milder (aOR 1.357 [95% CI 0.764-2.409], pinteraction=0.122).

**Conclusions:** Patients with LVO stroke of milder clinical severity have significantly better imaging markers of collateral circulation compared to high-severity patients. Despite high rates of thrombolytics, secondary transfer to thrombectomy-center was only associated with poorer 90-day functional outcomes in higher-severity patients. Failure of pre-hospital screening tools to detect lower-severity LVO patients for immediate pre-hospital bypass to a thrombectomy center may not deleteriously affect their outcome.

## Introduction

For patients with large vessel occlusion (LVO), endovascular thrombectomy (EVT) with or without intravenous thrombolytics dramatically improves functional post-stroke outcome.^1^ As EVT efficacy sharply diminishes with time,^2^ delays from secondary inter-hospital transfers for EVT (as opposed to direct EVT-center presentation) are associated with harm.^3,4^ Pre-hospital triage tools for selecting likely EVT candidates reduces need for secondary transfers, but there is considerable uncertainty in balancing tool sensitivity and specificity^5^ based on the accepted threshold of clinical severity that triggers a positive tool assessment.

Approximately 30% of patients with LVO have milder baseline severity^6^ and we hypothesized such patients have better collateral circulation that maintains blood flow above the symptomatic threshold and have slower infarct growth kinetics. Consequently, such patients may experience less harm from secondary transfer delays. This study therefore aimed to investigate the association of milder clinical severity with imaging markers of collateral adequacy and functional outcomes in patients needing secondary transfer.

## Materials and methods

### Patient inclusion

We performed a multicenter, retrospective cohort study using open-label stroke registry data from two major Australian metropolitan EVT centers (Royal Melbourne Hospital and Royal Adelaide Hospital) including consecutive patients who directly presented or underwent secondary transfer for EVT from 2016 to 2021, with complete 90-day follow-up data. We dichotomized baseline severity as milder (NIHSS <10) and severe (NIHSS ≥10) based on the likely threshold of LVO detection of commonly used triage tools using an earlier dataset.^6^ Data collection was approved by institutional ethics committees with waiver of consent. The study complies with STROBE observational cohort guidelines (https://www.strobe-statement.org/). Authors H.R. and H.Z. had full access to all data and are responsible for integrity and data analysis.

### Outcomes

To compare adequacy of collateral circulation between patients with milder and severe baseline severity, we used previously validated baseline CT-perfusion parameters^7^ obtained through RAPID processing software (iSchemaView, Inc.): 1) estimated ischemic core volume (cerebral blood volume <30%), 2) critically hypoperfused tissue (time-to-maximum >6 secs), 3) perfusion mismatch ratio (ratio of time-to-maximum >6 secs volume/cerebral blood volume <30% volume) and 4) hypointensity intensity ratio (ratio of time-to-maximum >10 secs volume/>6 secs volume). Functional outcomes were assessed at 90 days with the modified Rankin Scale (mRS) with a good outcome defined as mRS 0-2 or return to baseline mRS.

### Statistical analysis

Mann-Whitney U-test was used for continuous variables and Fisher exact test for categorical variables. The association of 90-day mRS in patients with milder versus severe deficits was investigated using logistic regression modelling adjusted for age, occlusion site and pre-morbid mRS. Transfer status by degree of deficit interactions were assessed by including respective multiplicative interaction terms in regression models. Sensitivity analyses were also conducted for those achieving good reperfusion (Thrombolysis In Cerebral Infarction grade 2b/3) and to only include internal carotid and first/second middle cerebral artery occlusions. Statistical analysis was performed using SPSS v27 (IBM Corp.) and Stata v17SE (StataCorp, TX, USA).

## Results

### Patient characteristics

Table 1 shows demographic and study outcomes. A total of 1210 LVO patients with complete 90-day mRS data were included, of which 273/1210 (22.6%) had milder baseline severity. Median onset-to-groin puncture time was 282 min (IQR 219-381) for secondarily transferred patients and 210 min (IQR 162-289) for patients presenting directly to a EVT-center (p<0.001). The vast majority (>90%) received intravenous thrombolysis, with median onset-to-needle 150min (IQR 115-199) for transfer and 148min (IQR 105-195) for direct presenting patients (p=0.058). Patients with milder baseline severity had lower rates of secondary transfer and longer onset-to-arterial access, while those with severe deficits had more proximal arterial occlusions at baseline.

**Table 1:**
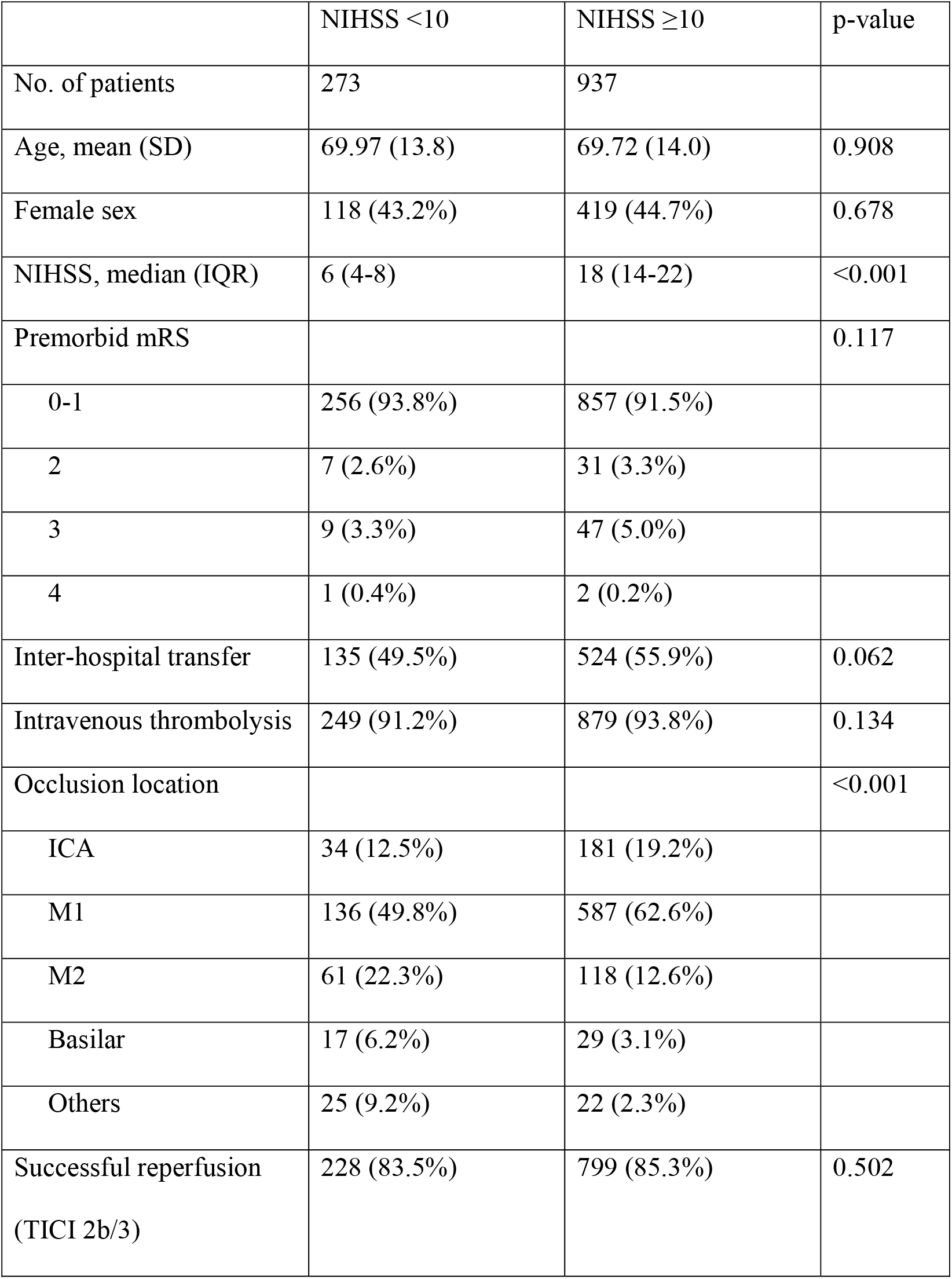
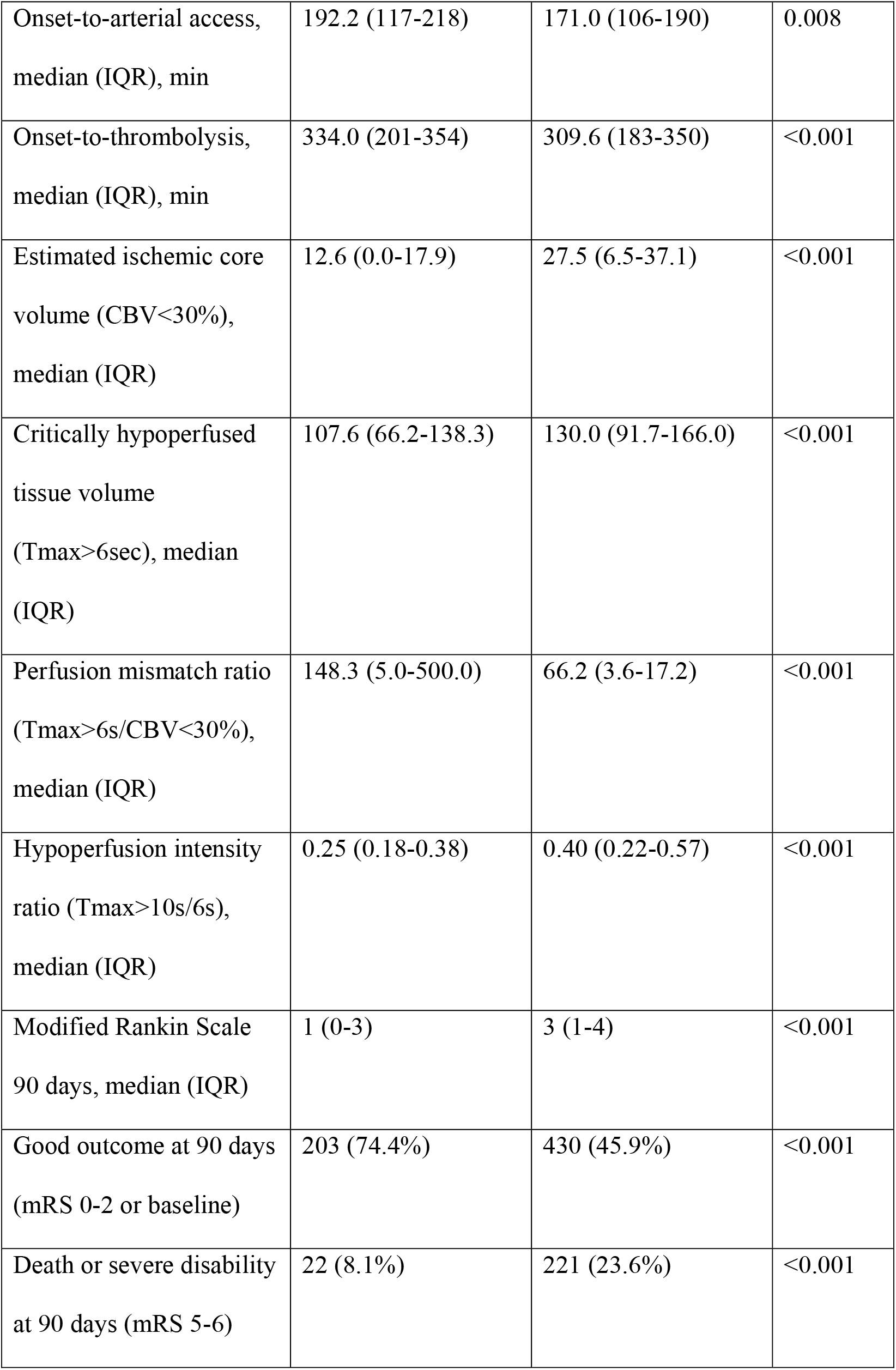
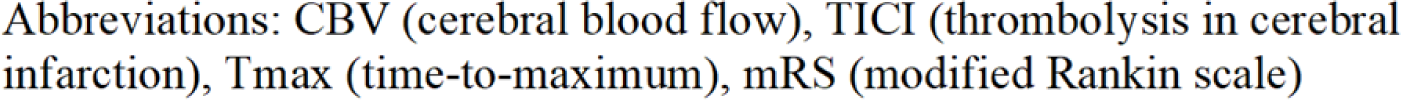
Baseline demographics

|  | NIHSS <10 | NIHSS ≥10 | p-value |
| --- | --- | --- | --- |
| No. of patients | 273 | 937 |  |
| Age, mean (SD) | 69.97 (13.8) | 69.72 (14.0) | 0.908 |
| Female sex | 118 (43.2%) | 419 (44.7%) | 0.678 |
| NIHSS, median (IQR) | 6 (4-8) | 18 (14-22) | <0.001 |
| Premorbid mRS |  |  | 0.117 |
| 0-1 | 256 (93.8%) | 857 (91.5%) |  |
| 2 | 7 (2.6%) | 31 (3.3%) |  |
| 3 | 9 (3.3%) | 47 (5.0%) |  |
| 4 | 1 (0.4%) | 2 (0.2%) |  |
| Inter-hospital transfer | 135 (49.5%) | 524 (55.9%) | 0.062 |
| Intravenous thrombolysis | 249 (91.2%) | 879 (93.8%) | 0.134 |
| Occlusion location |  |  | <0.001 |
| ICA | 34 (12.5%) | 181 (19.2%) |  |
| M1 | 136 (49.8%) | 587 (62.6%) |  |
| M2 | 61 (22.3%) | 118 (12.6%) |  |
| Basilar | 17 (6.2%) | 29 (3.1%) |  |
| Others | 25 (9.2%) | 22 (2.3%) |  |
| Successful reperfusion<br>(TICI 2b/3) | 228 (83.5%) | 799 (85.3%) | 0.502 |
| Onset-to-arterial access,<br>median (IQR), min | 192.2 (117-218) | 171.0 (106-190) | 0.008 |
| Onset-to-thrombolysis,<br>median (IQR), min | 334.0 (201-354) | 309.6 (183-350) | <0.001 |
| Estimated ischemic core<br>volume (CBV<30%),<br>median (IQR) | 12.6 (0.0-17.9) | 27.5 (6.5-37.1) | <0.001 |
| Critically hypoperfused<br>tissue volume<br>(Tmax>6sec), median<br>(IQR) | 107.6 (66.2-138.3) | 130.0 (91.7-166.0) | <0.001 |
| Perfusion mismatch ratio<br>(Tmax>6s/CBV<30%),<br>median (IQR) | 148.3 (5.0-500.0) | 66.2 (3.6-17.2) | <0.001 |
| Hypoperfusion intensity<br>ratio (Tmax>10s/6s),<br>median (IQR) | 0.25 (0.18-0.38) | 0.40 (0.22-0.57) | <0.001 |
| Modified Rankin Scale<br>90 days, median (IQR) | 1 (0-3) | 3 (1-4) | <0.001 |
| Good outcome at 90 days<br>(mRS 0-2 or baseline) | 203 (74.4%) | 430 (45.9%) | <0.001 |
| Death or severe disability<br>at 90 days (mRS 5-6) | 22 (8.1%) | 221 (23.6%) | <0.001 |

Abbreviations: CBV (cerebral blood flow), TICI (thrombolysis in cerebral infarction), Tmax (time-to-maximum), mRS (modified Rankin scale)

### Collateral circulation assessment

Baseline CT-perfusion data were processed for 436 patients. Patients with milder severity showed lower median estimated ischemic core volume (12.6 [IQR 0.0-17.9] vs 27.5 [IQR 6.5-37.1] mL, p<0.001) and lower median critically hypoperfused tissue volume (107.6 [IQR 66.2-138.3] vs 130 [IQR 91.7-166.0] mL, p<0.001). In addition, those with milder severity had greater median perfusion mismatch ratio (148.3 [IQR 5.0-500.0] vs 66.2 [IQR 3.6-17.2], p<0.001) and lower median hypoperfusion intensity ratio (0.25 [IQR 0.18-0.38] vs 0.40 [IQR 0.22-0.57], p<0.001).

### Functional outcome

Good outcome (mRS 0-2 or baseline) was achieved in 203(74.4%) patients with milder deficits and 430(45.9%) in patients with severe deficits (p<0.001). Table 2 shows the distribution of mRS scores for mild and severe deficit LVO patients depending on transfer status. Secondary transfer was associated with lower odds of good outcome for patients with severe deficits (aOR 0.76 [95% CI 0.58-1.00]) but not when deficits were milder (aOR 1.36 [95% CI 0.76-2.41], p_interaction_=0.12). Results were similar for secondary sensitivity analyses (see Online-only data supplement).

## Discussion

In this study, patients undergoing EVT with milder baseline clinical severity had lower proportions of very proximal arterial occlusions and significantly better CT-perfusion based imaging markers of collateral circulation compared with those with severe baseline deficits. Need for secondary inter-hospital transfer was associated with poorer outcomes for patients with severe baseline deficits, however this was not the case when clinical deficits were milder. However, statistical heterogeneity between severe and milder groups according to transfer status was not demonstrated.

The implications of our findings are that, although LVO patients with milder baseline severity are less likely to be identified by severity-based triage tools, they have better markers of collateral circulation which have been previously associated with slower infarct growth kinetics during secondary transfer for EVT.^8^ In contrast, patients with severe baseline deficits showed poorer outcomes from secondary transfer delays. Accordingly, lowering specificity to increase milder LVO stroke detection triage tool sensitivity may not be beneficial and may even be harmful, given the potential concerns of thrombolytic delay and EVT-center overburdening.

Up to 20% of patients with a visible vessel occlusion and initially mild symptoms subsequently deteriorate,^9^ possibly due to eventual collateral failure. As such, LVO patients with milder severity may still require secondary transfer for monitoring at an EVT-center. However, this could be completed with less urgency and better patient selection than immediate pre-hospital bypass.

The limitations of this study include its retrospective nature and relatively small sample of patients with milder severity, which may have limited interpretation of functional outcome data. Larger studies with greater precision are needed to confirm these findings. There may also have been selective referral of patients to EVT centers, creating potential bias in the cohort of secondarily transferred patients. Lastly, most of our included patients received intravenous thrombolysis, such that caution should be advised in extrapolation of results to thrombolysis-ineligible patients.

## Data Availability

Data is available on reasonable request from a suitably qualified investigator

## Non-standard Abbreviations and Acronyms

EVT: Endovascular thrombectomy
LVO: Large vessel occlusion
mRS: Modified Rankin Scale
NIHSS: National Institutes of Health Stroke Scale

## Acknowledgements

Nil

---

**Figure 1:**
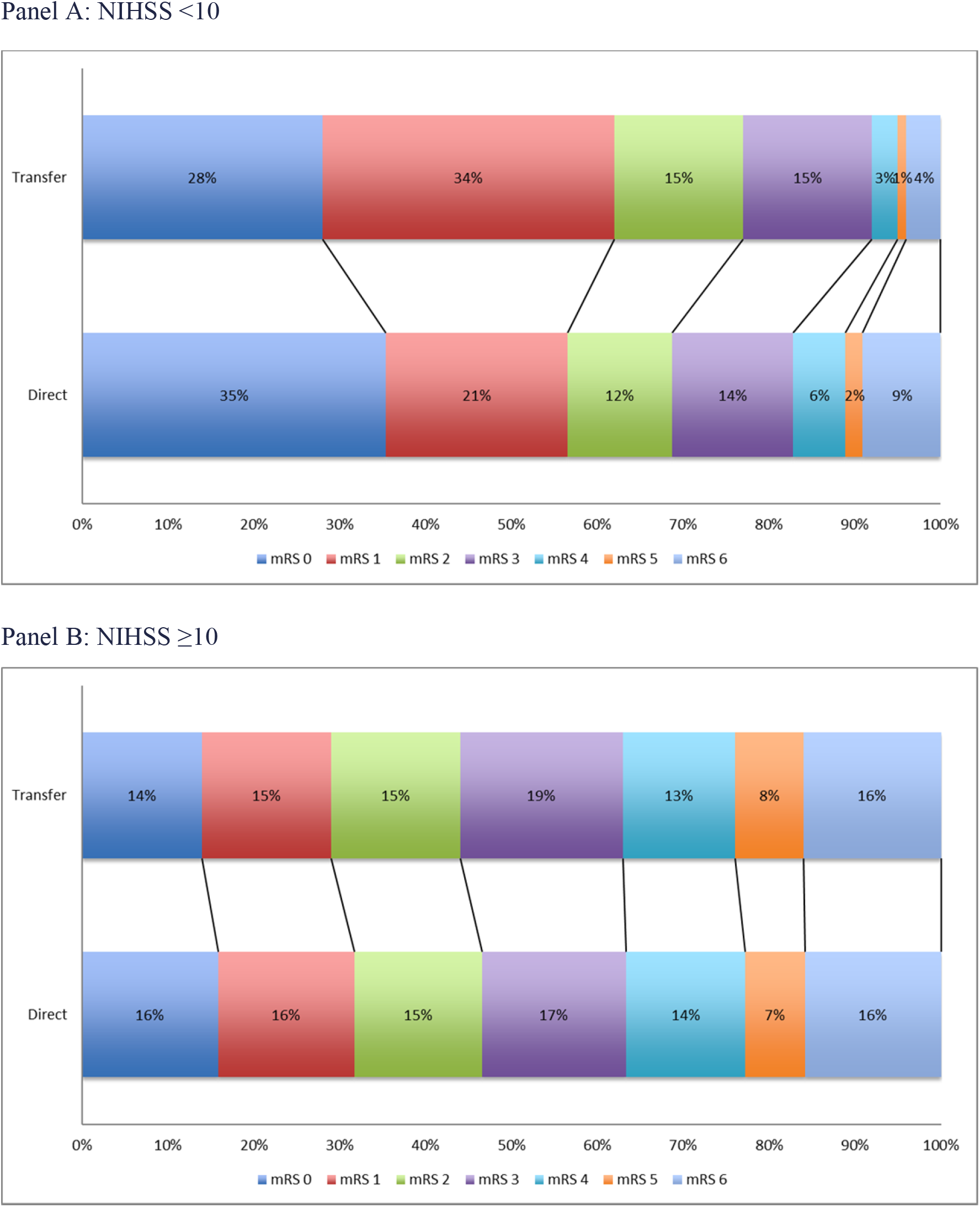
Association of transfer status with functional outcome in patients with differing baseline severities

